# Development of hybrid immunity during a period of high incidence of infections with Omicron subvariants: A prospective population based multi-region cohort study

**DOI:** 10.1101/2022.10.14.22281076

**Authors:** Anja Frei, Marco Kaufmann, Rebecca Amati, Audrey Butty Dettwiler, Viktor von Wyl, Anna Maria Annoni, Céline Pellaton, Giuseppe Pantaleo, Jan S. Fehr, Valérie D’Acremont, Murielle Bochud, Emiliano Albanese, Milo A. Puhan, the Corona Immunitas Research Group

## Abstract

**Background:** Seroprevalence and the proportion of people with neutralizing activity against SARS-CoV-2 variants was high in early 2022. Since it is unclear how immunity in the general population evolves, the aim of this study was to assess the development of functional and hybrid immunity in the general population during a period of high incidence of infections with Omicron variants.

**Methods:** This prospective population based multi-region cohort study is part of the Corona Immunitas research programme in Switzerland. In March 2022, we randomly selected individuals from the general population in southern (canton of Ticino) and north-eastern (canton of Zurich) Switzerland, who were assessed again in June/July 2022. We supplemented the June/July 2022 sample with a random sample from western Switzerland (canton of Vaud). We assessed SARS-CoV-2 specific IgG antibodies against spike and nucleocapsid proteins and the presence of SARS-CoV-2 neutralising antibodies against three variants (wildtype, Delta, Omicron).

**Findings:** In June/July 2022, seroprevalence was >98% in 2553 individuals from the general Swiss population. The proportion of individuals with neutralising antibodies against wildtype, Delta, and Omicron was 94.2%, 90.8%, and 84.9%, and at least 51% of the participants developed hybrid immunity. Individuals with hybrid immunity had, compared to those with only vaccine- or infection-induced immunity, highest levels of both, anti-spike IgG antibodies titres (4518 vs. 4304 vs. 269 WHO U/ml) and neutralisation capacity against wildtype (99.8% vs. 98% vs. 47.5%), Delta (99% vs. 92.2% vs. 38.7%), and Omicron (96.4% vs. 79.5% vs. 47.5%).

**Interpretation:** This first study on functional and hybrid immunity in the general population after Omicron waves showed that SARS-CoV-2 has become endemic. The high levels of antibodies and neutralization in the general populations support the emerging recommendations of some countries where booster vaccinations are still strongly recommended for vulnerable persons but less strongly recommended for individuals in the general population.

**Funding:** The Corona Immunitas research network is coordinated by the Swiss School of Public Health (SSPH+) and funded by fundraising of SSPH+ including funds of the Swiss Federal Office of Public Health and private funders (ethical guidelines for funding stated by SSPH+ were respected), by funds of the cantons of Switzerland (Vaud, Zurich, and Basel), and by institutional funds of the Universities.

**Study registration:** ISRCTN18181860

**Research in context:** *Evidence before this study:* We searched Pubmed, Medline, Scopus and Web of Knowledge, for primary population-based studies prospectively assessing infection-, vaccine-induced, and hybrid immunity and the respective neutralising activity of antibodies against SARS-CoV-2 and its variants of concern. We included articles published between 1 January and 28 September 2022, without language restrictions, and retrieved 540 publications after deduplication. None of the screened studies measured the prevalence of immune response and neutralisation capacity prospectively in population-based, representative samples accounting for type of acquired immunity. Evidence from five studies, all conducted in non-representative, convenience and relatively small samples (N<254), and/or in sub-populations (e.g., healthcare workers and children), shows that hybrid immunity confers higher immune protection and exhibits better neutralising capacity compared to vaccine- and infection-induced immunity. Furthermore, one of the screened studies highlights that antibodies developed by individuals with hybrid immunity show the slowest decline over a period of 10 months.

*Added value of this study:* We took advantage of an ongoing cohort study on anti-SARS-CoV-2 seroprevalence conducted in a representative sample of the general Swiss population (N=2553) using standard, previously validated methods, to measure changes over time in seroprevalence, neutralisation capacity against wildtype and variants of concerns of the virus (i.e., ACE2r-block), waning of antibodies, and new infections. This is the first study, conducted in the general population and during the pandemic phase characterized by very high incidence of Omicron infections, to assess the extent of hybrid immunity (51%) and neutralising antibodies against the wildtype (94.2%), Delta (90.8%), and Omicron variants (84.9%). Our findings show that individuals with hybrid immunity, compared to those with only vaccine- or infection-induced immunity, had the highest levels of both anti-spike IgG antibodies titres and neutralisation capacity against wildtype, Delta, and Omicron variants. We also found that, from March to June/July 2022, anti-spike IgG antibodies remained stable in the general population (>96%), while anti-nucleocapsid IgG antibodies fluctuated due to their fast waning (7.3% of participants’ anti-nucleocapsid IgG antibodies became undetectable) and the parallel spread of Omicron infections (18.6% of participants acquired anti-nucleocapsid IgG antibodies).

*Implications of all the available evidence:* By mid-2022, SARS-CoV-2 has become endemic, and a majority of individuals developed hybrid immunity with high levels of neutralisation against the wildtype, Delta, and Omicron variants of SARS-CoV-2. Combined with existing evidence, our results indicate that hybrid immunity confers higher levels of neutralising activity compared to both vaccine-induced and infection-induced immunity. This study extends findings on the immunological protection conferred by hybrid immunity from sub-populations to the general population. The high levels of antibodies and neutralization in the general populations support the emerging recommendations of some countries where booster vaccinations are still strongly recommended for vulnerable persons but less strongly recommended for individuals in the general population. Monitoring the prevalence, waning, and neutralising activity of antibodies against potential new variants of concern in populations remains crucial.

## Introduction

The WHO recently emphasized the key importance of continuous monitoring of immunity against SARS-CoV-2 and its variants of concern (VOC) in the general population to inform public health measures and vaccination strategies^1^. Early seroprevalence studies in spring 2020 showed that up to 10% of the general population had already developed antibodies against SARS-CoV-2 after the first wave in Europe and North America.^2–5^ Up to the point where vaccinations were approved in late 2020, seroprevalence increased to, on average, 25% as a consequence of SARS-CoV-2 infections but with great variations within and across countries.^1,6,7^ Following the introduction of vaccines, seroprevalence quickly increased to around 50% in the general population worldwide and to above 90% in high income countries.^6,8^ The concomitant reduction of the rate of severe disease courses of COVID-19 in vaccinated individuals provides strong support for the effectiveness of vaccines. However, although vaccines confer very high individual protection against COVID-19 symptoms, hospitalization and deaths, protection against new infections is partial, may decrease over time and with the emergence of new VOCs that can escape previously induced immunity.^9–11^

The rise of the highly infectious Omicron VOCs, in early 2022, caused many infections in fully vaccinated or boostered persons. This led to a high seroprevalence and functional immunity in the general population, as measured by neutralising activity of antibodies in serum.^12^ Functional immunity contributes to protection from severe courses of COVID-19 and is stronger if induced by both vaccinations and infections than either alone (i.e. hybrid immunity).^13–15^ To inform public health measures and further booster vaccine strategies, it is important to assess population levels of seroprevalence, and durability of functional and hybrid immunity developed during a time of high incidence of Omicron infections. The aim of this study was to assess the trajectory of anti-SARS-CoV-2 antibody titres, functional and hybrid immunity in the general population, and to compare such trajectories across age groups and three cantons representative of the three main regions in Switzerland.

## Methods

### Study design, sampling, and participants

This prospective population based multi-region cohort study is part of the Corona Immunitas research programme in Switzerland^16,17^, for which we had completed four phases of seroprevalence studies between April 2020 and October 2021 using a standardised protocol. The current study includes results from phases five and six, for which assessments were conducted between 1 March and 1 April 2022 and 30 May and 11 July, respectively.^12^

In phase five, we randomly selected individuals from the general population in southern (canton of Ticino) and north-eastern (canton of Zurich) Switzerland, who were assessed again in phase six. For cross-sectional analyses in phase six, we supplemented the southern and eastern Switzerland sample with a random sample from the general population in western Switzerland (canton of Vaud). These three Swiss cantons differ across demographic, socio-cultural, linguistic aspects and climate, all of which may impact on the dynamics of the pandemic.^18^ However, they are fairly representative for their language region (Italian, French, and German). The Swiss Federal Office of Statistics provided random samples of the general population in age-stratified (16–29, 30–44, 45–64 and ≥ 65 years) groups, separately for the cantons of Ticino, Vaud, and Zurich. We selected these groups after consultation with the Swiss Federal Office of Public Health to adequately account for the potential impact on seroprevalence of social behaviour, adherence to public health measures and vaccination uptake, all of which differ across these age groups.^19^ Given a test sensitivity of 97% and specificity of 99% of the seroprevalence test that we have used^20^, we deemed 200 participants for each stratum to provide precise enough estimates for an expected seroprevalence of 90% or more. The target sample size was thus 200 for each age stratum in the three cantons (i.e., total planned sample size of 2,400). Participation rate in phase five was 18.1% in Ticino (850 / 4687), 21.4% in Zurich (1044 / 4875), and 12.2% in Vaud (850 / 6963) in phase six. All phase five participants in Ticino and Zurich were invited to participate in the phase six blood sampling, of which 86.9% (739 / 850) in Ticino and 92.3% (964 / 1044) in Zurich decided to participate.

The study was registered (https://doi.org/10.1186/ISRCTN18181860) and approved by the ethics committees of the cantons of Zurich (BASEC Registration No 2020-01247), Ticino (BASEC Registration No 2020-01514), and Vaud (BASEC No 2020-00887). All participants provided written informed consent before participation.

### Data collection

We invited participants to in-person study visits at a healthcare facility to provide a blood sample. People who were not able or willing to travel were offered at-home visits. For each participant, trained personnel collected venous blood samples, according to clinical standards and COVID-19 hygiene measures. Before the first study visit, all participants completed a baseline questionnaire including information regarding socio-demographics, vaccinations, SARS-CoV-2 infections, hospital and intensive care unit (ICU) admissions, and symptoms in case of infections and past medical history, using the secure, web-based Research Electronic Data Capture platform (REDCap) for data collection and management.^21,22^ They also had the possibility to fill in the questionnaire on a paper/pencil version. Participants from the cantons of Ticino and Zurich who were recruited in phase five were invited for a second study visit and blood sampling in phase six, three to four months later. Before this second study visit, participants filled in another questionnaire targeting the time between the first and second blood sampling including questions on new self-reported SARS-CoV-2 infections, symptoms, and vaccinations.

### Laboratory assays for SARS-CoV-2 antibodies and neutralising capacity against SARS-CoV-2 variants

We assessed SARS-CoV-2 specific antibodies against the spike and nucleocapsid proteins using Sensitive Anti-SARS-CoV-2 Spike Trimer Immunoglobulin Serological (SenASTrIS), a Luminex binding assay.^20^ The assay measures binding of IgG antibodies to the trimeric SARS-CoV-2 spike and the nucleocapsid proteins. The test has a high specificity (99%) and sensitivity (97%) and has been validated in samples of the general population and in specific subgroups.^20^ The semi-quantitative mean fluorescence intensity (MFI) values have been dichotomised at the cut-off value of 6 (<6 not detectable, ≥6 detectable) and the height of the antibody response categorised (< 6 not detectable, ≥6 and <12 low, ≥12 and <40 middle, ≥40 high). Furthermore, the MFI have been translated to the U/mL scale as measured by the Elecsys Anti-SARS-CoV-2 immunoassay by Roche.^12^ We also assessed the presence of SARS-CoV-2 neutralising antibodies against three variants (wildtype, Delta, and Omicron) that were or are still dominant in Switzerland in 2022 using a cell- and virus-free assay.^23^ This assay measures the proportion of antibodies that block the interaction of the angiotensin-converting enzyme 2 receptor (ACE2r) with the receptor-binding domain of the trimer spike protein of the wildtype and variants of concern. Neutralisation has been determined to occur with a value above the cut-off value of 50.

### Outcome definition

We defined seropositivity based on the presence of anti-spike IgG antibodies according to the threshold of SenASTrIS test positivity with mean MFI ≥ 6, and neutralisation capacity based on the cut-off value of 50. Functional immunity was defined based on neutralisation capacity of the cell- and virus free assay above the threshold value of 50. This was determined independently for each variant spike (wildtype, Delta, Omicron). Lastly, source of immune status was defined based on SARS-CoV-2 vaccination status (self-reported) and SARS-CoV-2 infection, determined as seropositivity for anti-nucleocapsid IgG (MFI≥ 6), report of a positive PCR or rapid antigen test or presence of anti-spike IgG antibodies in the absence of a SARS-CoV-2 vaccination. We categorised immune status as follows: immune naïve (i.e., no detectable antibodies and no reported infection and SARS-CoV-2 vaccination), vaccine-induced only, infection-induced only, or hybrid immunity (SARS-CoV-2 vaccination and infection).

### Statistical analysis

We used medians and interquartile ranges or absolute and relative numbers for the descriptive analyses. We calculated seroprevalence using a Bayesian logistic regression model accounting for the accuracy characteristics of the serological test and applied post-stratification weights based on the target population demographic structure.^2^ We conducted all analyses in R, version 4.2.1.

We determined the proportion of individuals in whom anti-spike IgG antibodies remained negative or positive (i.e., unchanged) or changed from negative to positive or positive to negative.

## Results

Between 30 May 2022 and 11 July 2022 (phase six of the Corona Immunitas research programme), we assessed 2553 cohort participants, 739 from Ticino, 850 from Vaud, and 964 from Zurich (Table 1). Median age of the participants of the three cantons was respectively 49 (interquartile range [IQR] 35-64), 55 (IQR 39-69), and 52 years (IQR 35-66). Female participants were slightly overrepresented with proportions of 58.1%, 55.9%, and 54.4%, respectively in the three cantons. Most participants had received at least one dose of a SARS-CoV-2 vaccine (89.9%, 91.1%, and 93.6%). Around half of the study sample reported to have been infected recently, likely in 2022 (53%, 48.5%, 51.8%).

**Table 1:**
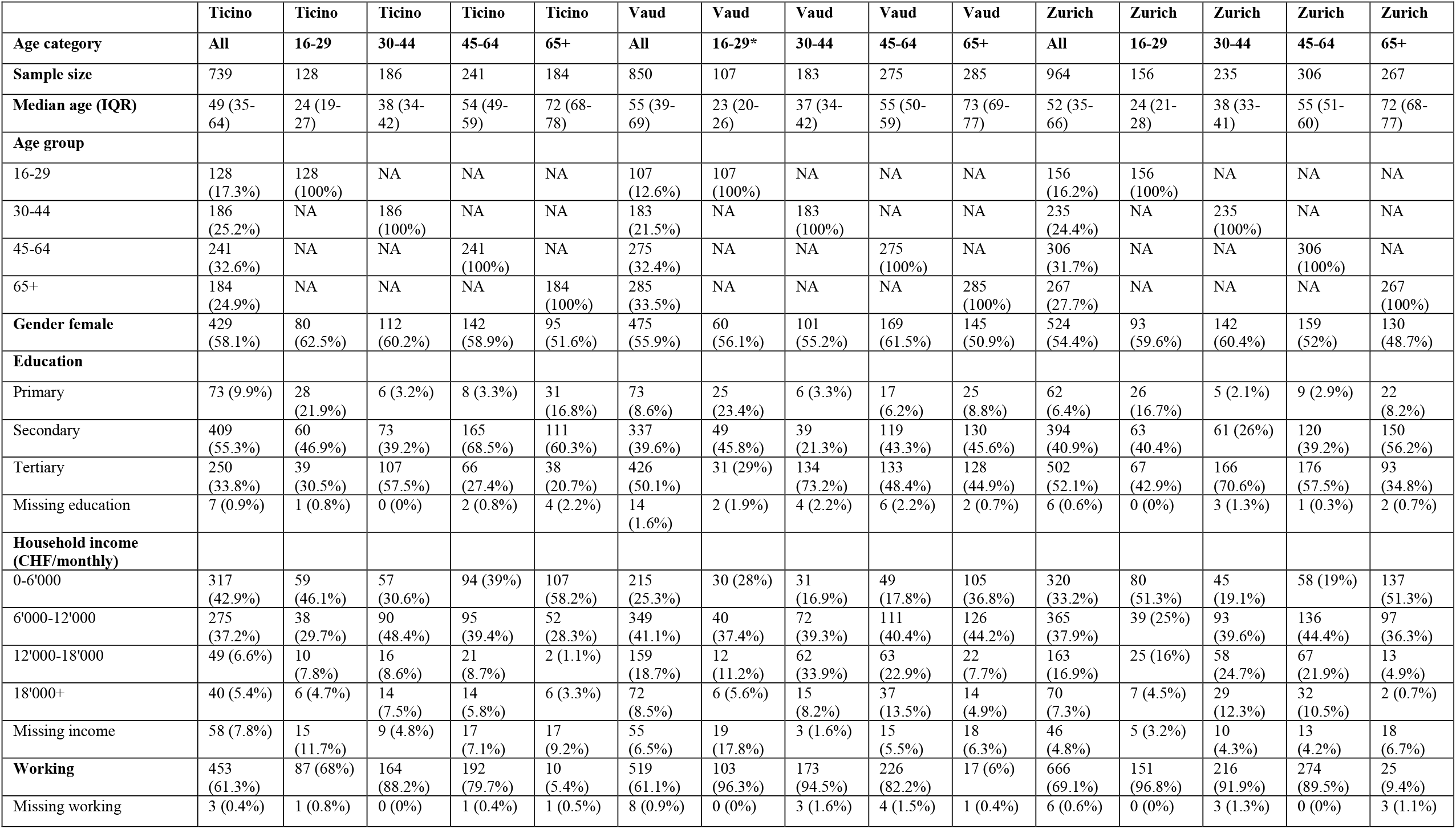
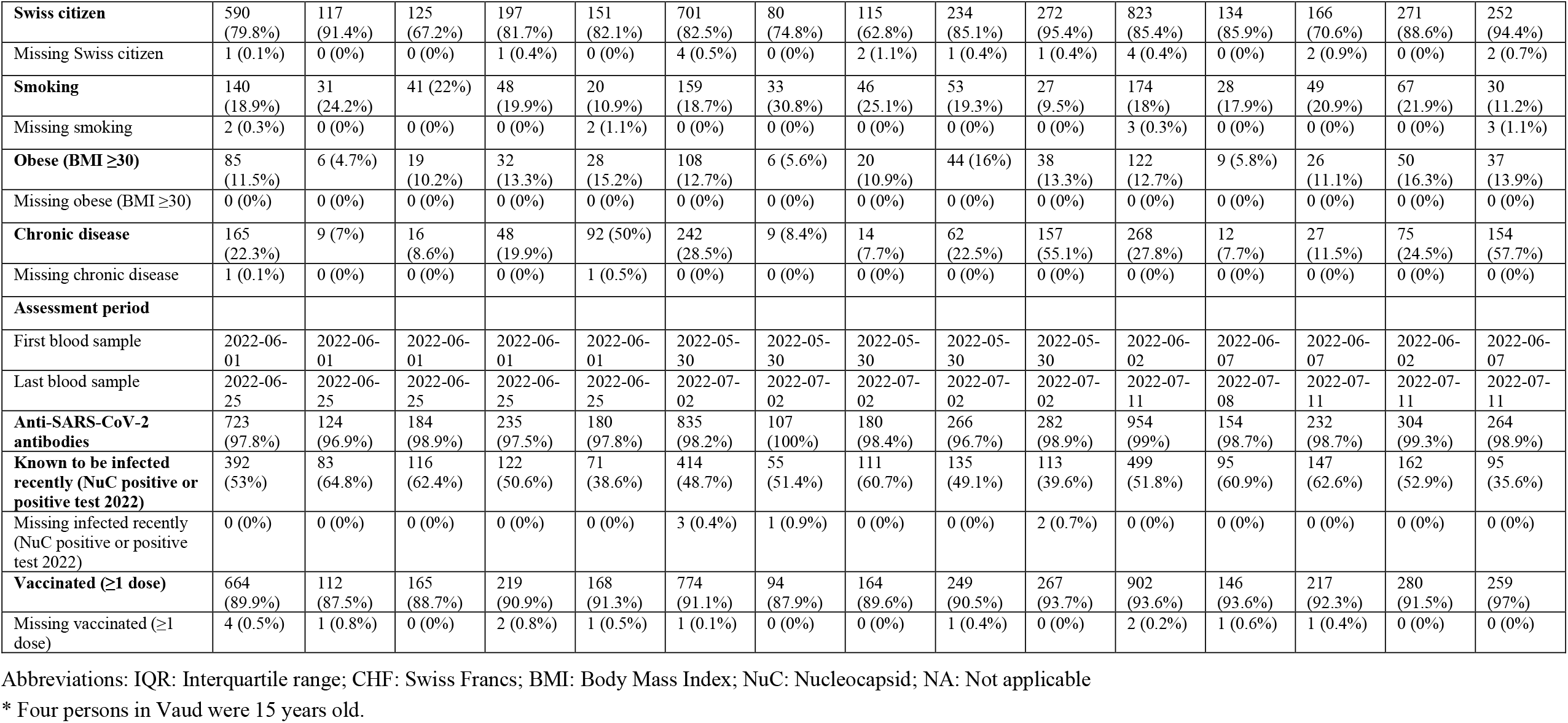
Characteristics of the sample, stratified by canton and age group. Ticino, Vaud, and Zurich, Switzerland, June-July 2022 (n=2553).

By June/July 2022, seroprevalence for Ticino, Vaud, and Zurich participants was estimated at 98.3% (95% confidence interval [CI] 96.9–99.3%), 98.4% (95% CI 97.3–99.3%), and 98.9% (95% CI 98–99.5%, Table 2). Anti-spike IgG antibodies were high across cantons and age-groups (>90%). The proportion of individuals whose antibodies showed neutralisation (ACE2r-blocking) was high against the wildtype (93.1% for Ticino, 93.9% for Vaud, 95.4% for Zurich) and Delta (90.7%, 91.8%, 90%), and only slightly lower for the Omicron (84.3%, 86.9%, 83.6%) variant of SARS-CoV-2, with no evident patterns across age groups and study sites.

**Table 2:**
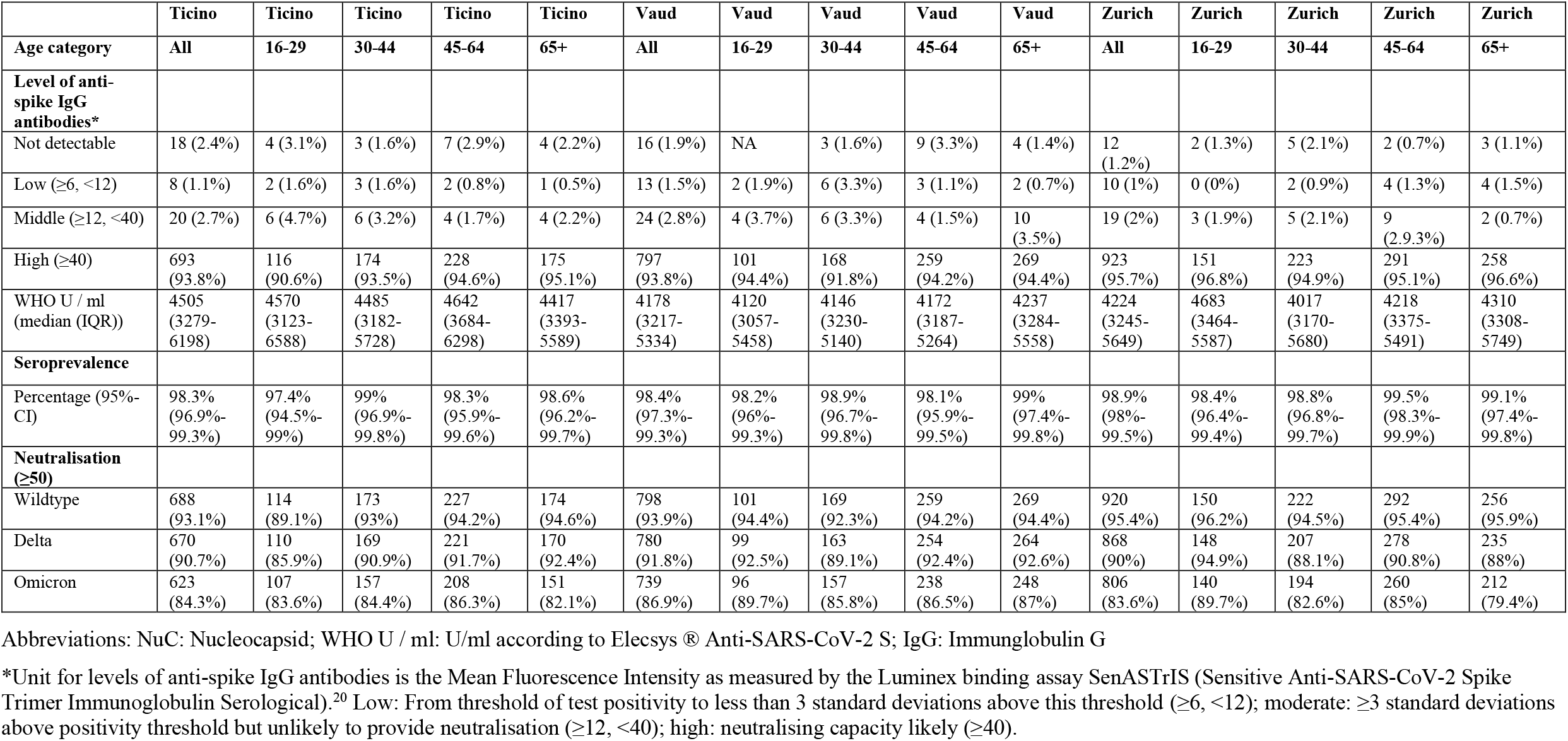
Prevalence of SARS-CoV-2 IgG antibodies and ACE2r-blocking (neutralising capacity) as measured by a virus-free assay, stratified by canton and age group. Ticino, Vaud, and Zurich, Switzerland, June-July 2022, (n=2553).

|  | Ticino | Ticino | Ticino | Ticino | Ticino | Vaud | Vaud | Vaud | Vaud | Vaud | Zurich | Zurich | Zurich | Zurich | Zurich |
| --- | --- | --- | --- | --- | --- | --- | --- | --- | --- | --- | --- | --- | --- | --- | --- |
| Age category | All | 16-29 | 30-44 | 45-64 | 65+ | All | 16-29 | 30-44 | 45-64 | 65+ | All | 16-29 | 30-44 | 45-64 | 65+ |
| Level of anti-spike IgG antibodies* |  |  |  |  |  |  |  |  |  |  |  |  |  |  |  |
| Not detectable | 18 (2.4%) | 4 (3.1%) | 3 (1.6%) | 7 (2.9%) | 4 (2.2%) | 16 (1.9%) | NA | 3 (1.6%) | 9 (3.3%) | 4 (1.4%) | 12 (1.2%) | 2 (1.3%) | 5 (2.1%) | 2 (0.7%) | 3 (1.1%) |
| Low ($\geq 6$, $< 12$) | 8 (1.1%) | 2 (1.6%) | 3 (1.6%) | 2 (0.8%) | 1 (0.5%) | 13 (1.5%) | 2 (1.9%) | 6 (3.3%) | 3 (1.1%) | 2 (0.7%) | 10 (1%) | 0 (0%) | 2 (0.9%) | 4 (1.3%) | 4 (1.5%) |
| Middle ($\geq 12$, $< 40$) | 20 (2.7%) | 6 (4.7%) | 6 (3.2%) | 4 (1.7%) | 4 (2.2%) | 24 (2.8%) | 4 (3.7%) | 6 (3.3%) | 4 (1.5%) | 10 (3.5%) | 19 (2%) | 3 (1.9%) | 5 (2.1%) | 9 (2.9.3%) | 2 (0.7%) |
| High ($\geq 40$) | 693 (93.8%) | 116 (90.6%) | 174 (93.5%) | 228 (94.6%) | 175 (95.1%) | 797 (93.8%) | 101 (94.4%) | 168 (91.8%) | 259 (94.2%) | 269 (94.4%) | 923 (95.7%) | 151 (96.8%) | 223 (94.9%) | 291 (95.1%) | 258 (96.6%) |
| WHO U / ml (median (IQR)) | 4505 (3279-6198) | 4570 (3123-6588) | 4485 (3182-5728) | 4642 (3684-6298) | 4417 (3393-5589) | 4178 (3217-5334) | 4120 (3057-5458) | 4146 (3230-5140) | 4172 (3187-5264) | 4237 (3284-5558) | 4224 (3245-5649) | 4683 (3464-5587) | 4017 (3170-5680) | 4218 (3375-5491) | 4310 (3308-5749) |
| Seroprevalence |  |  |  |  |  |  |  |  |  |  |  |  |  |  |  |
| Percentage (95%-CI) | 98.3% (96.9%-99.3%) | 97.4% (94.5%-99%) | 99% (96.9%-99.8%) | 98.3% (95.9%-99.6%) | 98.6% (96.2%-99.7%) | 98.4% (97.3%-99.3%) | 98.2% (96%-99.3%) | 98.9% (96.7%-99.8%) | 98.1% (95.9%-99.5%) | 99% (97.4%-99.8%) | 98.9% (98%-99.5%) | 98.4% (96.4%-99.4%) | 98.8% (96.8%-99.7%) | 99.5% (98.3%-99.9%) | 99.1% (97.4%-99.8%) |
| Neutralisation ($\geq 50$) | | | | | | | | | | | | | | | |
| Wildtype | 688 (93.1%) | 114 (89.1%) | 173 (93%) | 227 (94.2%) | 174 (94.6%) | 798 (93.9%) | 101 (94.4%) | 169 (92.3%) | 259 (94.2%) | 269 (94.4%) | 920 (95.4%) | 150 (96.2%) | 222 (94.5%) | 292 (95.4%) | 256 (95.9%) |
| Delta | 670 (90.7%) | 110 (85.9%) | 169 (90.9%) | 221 (91.7%) | 170 (92.4%) | 780 (91.8%) | 99 (92.5%) | 163 (89.1%) | 254 (92.4%) | 264 (92.6%) | 868 (90%) | 148 (94.9%) | 207 (88.1%) | 278 (90.8%) | 235 (88%) |
| Omicron | 623 (84.3%) | 107 (83.6%) | 157 (84.4%) | 208 (86.3%) | 151 (82.1%) | 739 (86.9%) | 96 (89.7%) | 157 (85.8%) | 238 (86.5%) | 248 (87%) | 806 (83.6%) | 140 (89.7%) | 194 (82.6%) | 260 (85%) | 212 (79.4%) |

Abbreviations: NuC: Nucleocapsid; WHO U / ml: U/ml according to Elecsys<sup>®</sup> Anti-SARS-CoV-2 S; IgG: Immunglobulin G
\*Unit for levels of anti-spike IgG antibodies is the Mean Fluorescence Intensity as measured by the Luminex binding assay SenASTrIS (Sensitive Anti-SARS-CoV-2 Spike Trimer Immunoglobulin Serological).<sup>20</sup> Low: From threshold of test positivity to less than 3 standard deviations above this threshold ( $\geq 6$ , $< 12$ ); moderate: $\geq 3$ standard deviations above positivity threshold but unlikely to provide neutralisation ( $\geq 12$ , $< 40$ ); high: neutralising capacity likely ( $\geq 40$ ).

From March 2022 to June/July 2022, the proportion of participants from Ticino and Zurich with detectable anti-spike IgG antibodies remained stable (>96% across age groups), only in seven participants the anti-spike IgG decreased below the threshold, all of whom were unvaccinated and had become infected in 2022, most likely with the Omicron variant. In contrast, the fluctuation in anti-nucleocapsid IgG antibodies reflected both quick waning of anti-nucleocapsid antibodies (from positive to negative: 7.3%) as well as substantial infection activity with the Omicron variant in spring 2022 in Switzerland (from negative to positive: 18.6%). The neutralisation capacity against the wildtype, Delta, and Omicron variant remained stable (from positive to positive: 93.1%, 88.5%, and 80%, respectively), with little variation across age groups (Table 3 [pooled results] and Supplementary Table S1 [results stratified by canton]). There was a higher loss of neutralisation capacity (from positive to negative) observed for Omicron with 8.6% (wildtype 1.2%, Delta 4.2%), while on the population level only little changed with respect to newly obtained neutralisation capacity (form negative to positive: wildtype 1.3%, Delta 1.8%, Omicron 3.9%).

**Table 3:** Trajectories from March 2022 to June/July 2022 of SARS-CoV-2 IgG antibodies and ACE2r-blocking (neutralising capacity) as measured by a virus-free assay, in participants from Ticino and Zurich, Switzerland, pooled, stratified by age group (n=1702*).

|  | All | 16-29 | 30-44 | 45-64 | 65+ |
| --- | --- | --- | --- | --- | --- |
| **Anti-spike IgG antibodies** |  |  |  |  |  |
| MFI change - Median (IQR)** | 1776 (568 to 3203) | 1984 (670 to 3455) | 1626 (437 to 3196) | 1806 (684 to 3233) | 1807 (535 to 3011) |
| Negative - Negative | 23 (1.4%) | 2 (0.7%) | 5 (1.2%) | 9 (1.6%) | 7 (1.6%) |
| Negative - Positive | 15 (0.9%) | 4 (1.4%) | 3 (0.7%) | 4 (0.7%) | 4 (0.9%) |
| Positive - Negative | 7 (0.4%) | 4 (1.4%) | 3 (0.7%) | 0 (0%) | 0 (0%) |
| Positive - Positive | 1657 (97.4%) | 274 (96.5%) | 410 (97.4%) | 534 (97.6%) | 439 (97.6%) |
| **Anti-NuC IgG antibodies#** |  |  |  |  |  |
| Negative - Negative | 971 (57.1%) | 138 (48.6%) | 211 (50.1%) | 315 (57.6%) | 307 (68.2%) |
| Negative - Positive | 316 (18.6%) | 54 (19%) | 82 (19.5%) | 99 (18.1%) | 81 (18%) |
| Positive - Negative | 125 (7.3%) | 33 (11.6%) | 38 (9%) | 40 (7.3%) | 14 (3.1%) |
| Positive - Positive | 290 (17%) | 59 (20.8%) | 90 (21.4%) | 93 (17%) | 48 (10.7%) |
| **Neutralisation Wildtype##** |  |  |  |  |  |
| Negative - Negative | 75 (4.4%) | 17 (6%) | 20 (4.8%) | 23 (4.2%) | 15 (3.3%) |
| Negative - Positive | 22 (1.3%) | 3 (1.1%) | 7 (1.7%) | 8 (1.5%) | 4 (0.9%) |
| Positive - Negative | 20 (1.2%) | 3 (1.1%) | 6 (1.4%) | 5 (0.9%) | 6 (1.3%) |
| Positive – Positive | 1585 (93.1%) | 261 (91.9%) | 388 (92.2%) | 511 (93.4%) | 425 (94.4%) |
| **Neutralisation Delta##** |  |  |  |  |  |
| Negative - Negative | 94 (5.5%) | 19 (6.7%) | 22 (5.2%) | 28 (5.1%) | 25 (5.6%) |
| Negative - Positive | 30 (1.8%) | 4 (1.4%) | 7 (1.7%) | 11 (2%) | 8 (1.8%) |
| Positive - Negative | 71 (4.2%) | 7 (2.5%) | 23 (5.5%) | 20 (3.7%) | 21 (4.7%) |
| Positive – Positive | 1507 (88.5%) | 254 (89.4%) | 369 (87.6%) | 488 (89.2%) | 396 (88%) |
| **Neutralisation Omicron##** |  |  |  |  |  |
| Negative - Negative | 128 (7.5%) | 21 (7.4%) | 33 (7.8%) | 37 (6.8%) | 37 (8.2%) |
| Negative - Positive | 66 (3.9%) | 8 (2.8%) | 15 (3.6%) | 25 (4.6%) | 18 (4%) |
| Positive - Negative | 146 (8.6%) | 16 (5.6%) | 37 (8.8%) | 42 (7.7%) | 51 (11.3%) |
| Positive - Positive | 1362 (80%) | 239 (84.2%) | 336 (79.8%) | 443 (81%) | 344 (76.4%) |

Abbreviations: NuC: Nucleocapsid; WHO U / ml: U/ml according to Elecsys ® Anti-SARS-CoV-2 S; IgG: Immunglobulin G
\*For one participant from Ticino no results for phase five (March 2022) is available.
\*\*Unit for levels of anti-spike IgG antibodies is the Mean Fluorescence Intensity as measured by the Luminex binding assay SenASTrIS (Sensitive Anti-SARS-CoV-2 Spike Trimer Immunoglobulin Serological)<sup>20</sup>)
#Seropositivity is defined based on the presence of anti-spike IgG antibodies according to the threshold of SenASTrIS test positivity with mean MFI $\geq 6$ .

In June/July 2022, the proportion with high levels of anti-spike IgG antibodies was more than double in vaccinated individuals and persons with a hybrid immunity compared to individuals with an infection only (high anti-spike IgG antibodies 99% and 99.8% vs. 45.9%; Table 4 [pooled results] and Supplementary Table S2 [results stratified by canton]). Such large differences were also observed for neutralisation capacity (wildtype 98% and 99.8% vs. 47.5%; Delta 92.2% and 99% vs. 38.7%; Omicron 79.5% and 96.4% vs. 47.5%). Neutralisation against Delta and Omicron was highest in participants with hybrid immunity, followed by those who have only been vaccinated and much lower in those with infection only. Compared to March 2022 (phase five), hybrid immunity in participants from Ticino and Zurich increased from 35.8% to 50.6% by June/July 2022 (phase six), reflecting the high incidence with Omicron infections since spring 2022.

**Table 4:**
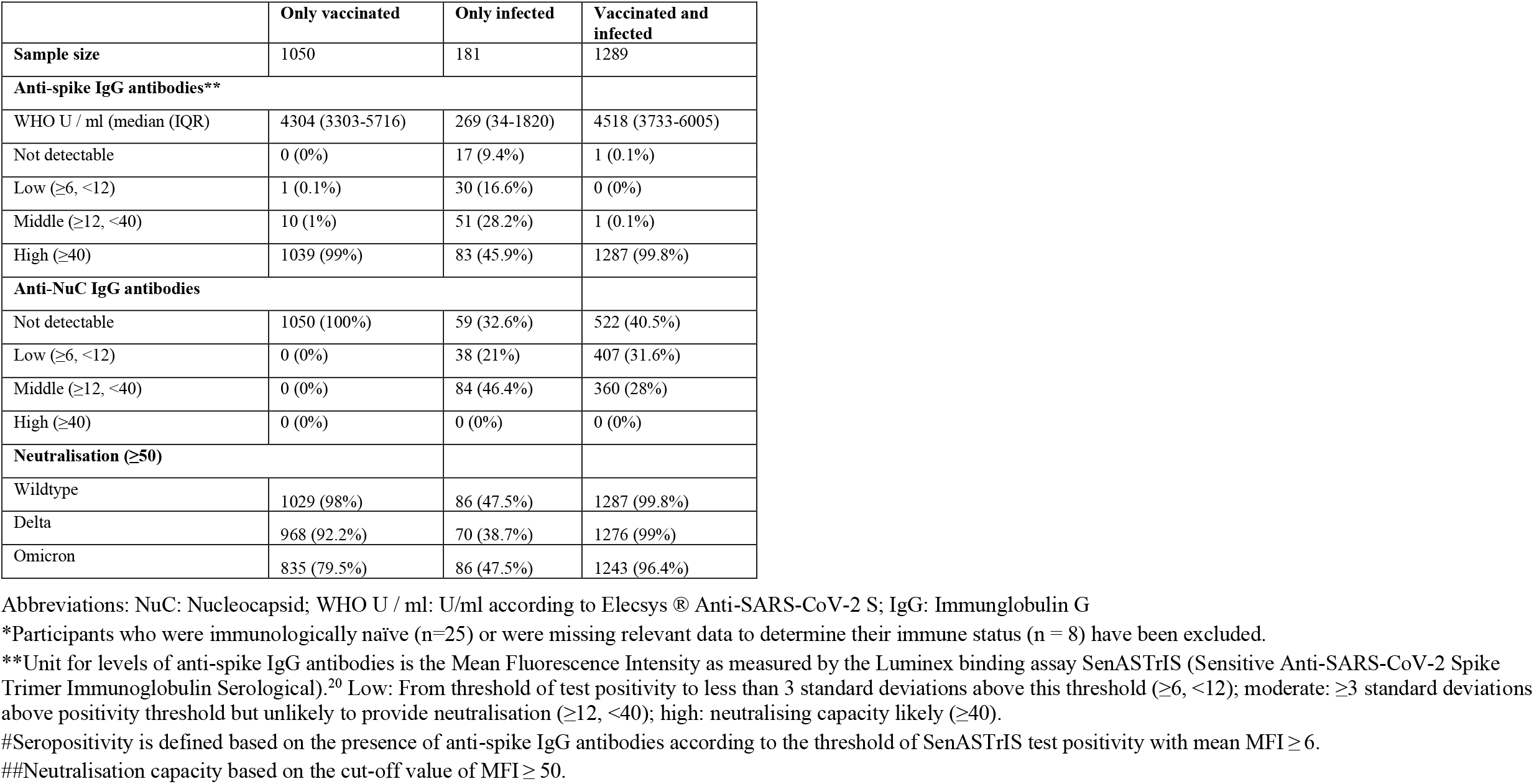
Prevalence of SARS-CoV-2 IgG antibodies and ACE2r-blocking (neutralising capacity) as measured by a virus-free assay, stratified by vaccination and infection status of participants, Ticino, Vaud, and Zurich, Switzerland, pooled, June/July 2022, (n=2520*).

## Discussion

Our population-based cohort study showed that not only seroprevalence but also antibody titres were very high in the general population in Switzerland by June/July 2022, without notable differences across cantons, age, and sex. At least 51% of study participants developed hybrid immunity and among those, more than 96% had neutralizing antibodies against the wildtype, Delta, and Omicron variants. In participants who received vaccination but were not infected previously, the proportion with neutralising antibodies was lower, in particular against Omicron. The 7% of participants with only an infection-induced immunity had about 15 times lower antibody titres, and less than 50% of them showed neutralizing antibodies. These findings are in line with previous studies, conducted in small and non-population-based samples, showing that hybrid immunity can confer to antibodies a better neutralisation capacity.^24–30^

This study is the first to demonstrate the extent of hybrid immunity and neutralisation capacity in the general population in 2022. It is likely that we underestimated hybrid immunity and that its prevalence is higher than 51%, as anti-nucleocapsid antibodies wane quickly and we likely missed some infections that occurred before 2022. Self-reports of infections compensate only to some extent for the low to moderate sensitivity of anti-nucleocapsid assays beyond 6 months of infection because many infections are mild or asymptomatic. A large study from Israel in 2021 showed that hybrid immunity provided stronger protection than vaccination and infection alone.^15^ While the proportion of persons with hybrid immunity was not reported, the observation time for persons with infection and vaccination up to (re) infection or censoring was smaller compared to those only vaccinated or only infected, implying a very low prevalence of hybrid immunity back in 2021.

Our results have implications for vaccination strategies. Recommendations for primary series and booster vaccination need to consider the effectiveness and safety of vaccines as well as the epidemiologic and societal context.^31^ Seroprevalence is only a rough proxy marker of immunity in the population since seropositive persons have a wide range of antibody titres and neutralising capacity against SARS-CoV-2 VOCs as a consequence of infection only, vaccination only or both infection and vaccination as this study and other studies showed.^13,14^ Therefore, information on the proportion of persons in the general population with neutralising capacity and hybrid immunity provides more solid guidance. The Swiss Federal Vaccination Commission recently released finely granulated recommendations for booster and primary series vaccinations based on the best available international evidence on the effectiveness and safety of bivalent or other booster vaccines, and based on the results of Corona Immunitas presented here. While the Commission issued a strong recommendation for a second booster for people above 64 years of age, for those with chronic conditions and pregnant women, the recommendation was moderately strong for health care staff and formal and informal caregivers, and only weak for the general population between 16 and 64 years of age. In addition, they recommended only one primary series dose for unvaccinated persons since most of them have had a SARS-CoV-2 infection (>90% according to the results presented here). These recommendations considered the high seroprevalence in Switzerland and the high proportion of persons with hybrid immunity and neutralising capacity, and include considerations on the optimal timing for the next booster campaign in autumn/winter 2022. The Canadian authorities issued similar recommendations for booster vaccines while population-based data on immunity in the population were not available to the extent and level of detail presented here.^32^

Strengths of our study include the prospective, population-based cohort study design, coverage of the three main language and cultural regions of a country, the well-established methods of the Corona Immunitas research programme, the large sample size, and the use of previously validated serological tests and neutralising antibodies.^20,23^ In addition, retention of participants since March 2022 was high. Limitations include the modest participation rate, as it is commonly the case in population-based studies, and the lack of measures of cellular immunity, which is not feasible to test in large population-based studies. An additional limitation is that recent infections may have been overlooked because of waning of anti-nucleocapsid antibodies and low participants’ awareness and reporting of recent infections. This likely led to an underestimation of hybrid immunity in our sample.

## Conclusion

This prospective population-based cohort study with 2553 participants showed that seroprevalence remained very high in Switzerland in 2022, without differences across cantons and age groups. Antibody titres increased, and the majority of participants developed hybrid immunity with very high levels of neutralisation against the wildtype, Delta, and Omicron variants of SARS-CoV-2. Individuals with immunity only from infection had 15 times lower antibody titres, and less than half of them showed neutralisation. Our results support the emerging recommendations of some countries where booster vaccinations are still strongly recommended for vulnerable persons but less strongly recommended for individuals in the general population.

**Figure 1.**
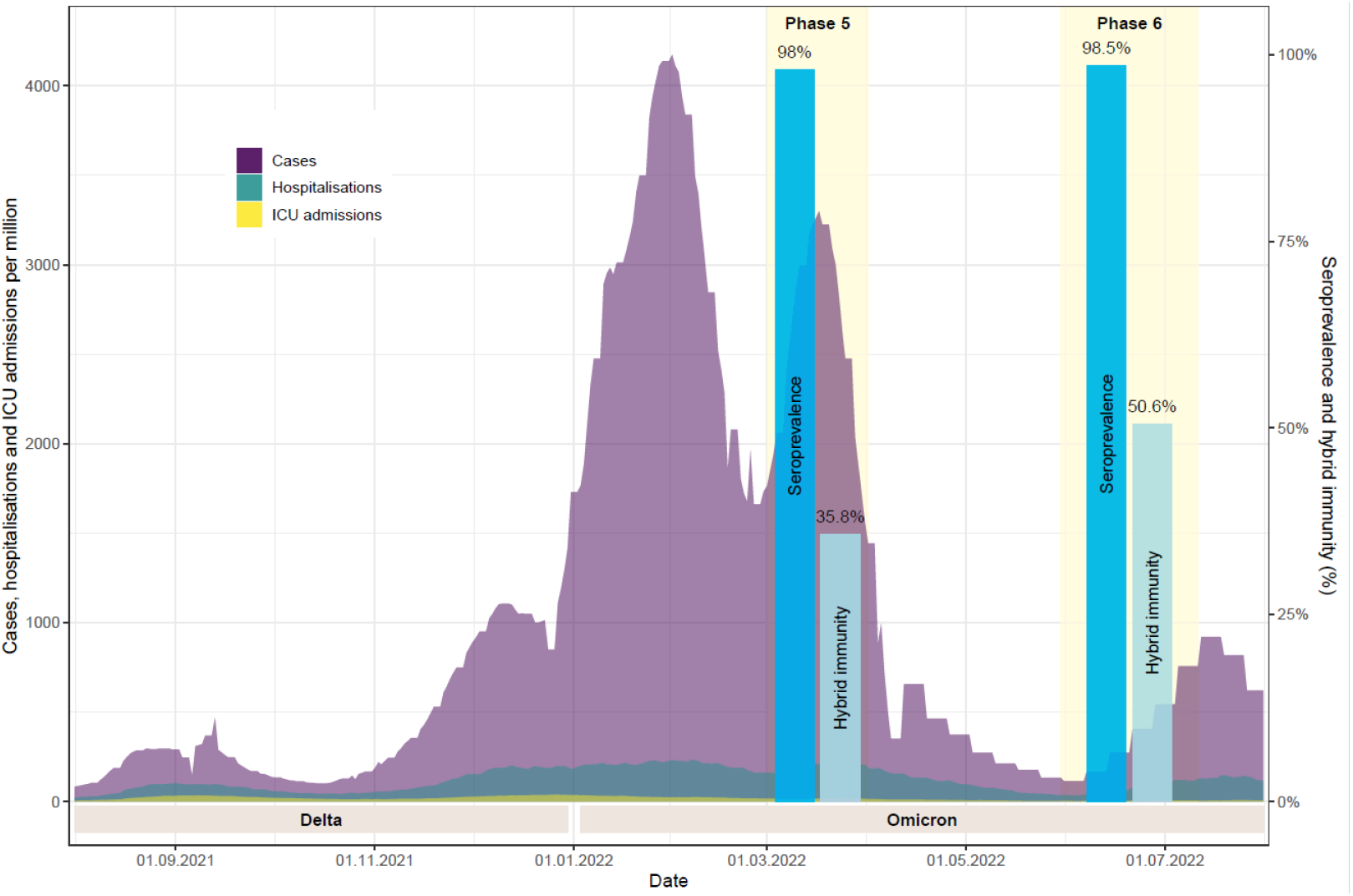
Seroprevalence and hybrid immunity in phases five and six of Corona Immunitas, Switzerland, in relation to the evolution of the pandemic, August 2021–August 2022. The evolution of the pandemic is visualised by the number of confirmed SARS-CoV-2 cases (in purple), hospitalisations (turquois), and intensive care unit (ICU) admissions in Switzerland between August 2021 and August 2022 (retrieved from: https://ourworldindata.org/coronavirus).^33^ The results regarding seroprevalence and proportion of participants with hybrid immunity are visualised in dark and light blue bars, respectively, for phases five (March 2022, n=1894) and six (June/July 2022, n=1702) of Corona Immunitas, cantons Ticino and Zurich, Switzerland.

## Data Availability

All data produced in the present study are available upon reasonable request to the authors after publication of this article

## Author contribution

MAP, EA, JF, AF, MK, MB, VDA, and RA conceptualized and designed this study. AF, RA, ABD, MK, VVW, and AMA contributed to the acquisition of the data. MK conducted the statistical analyses and drafted the figure; CP and GP contributed to the laboratory analyses; RA, AMA, and EA conducted the systematic literature research. All authors had full access to the data and contributed to the interpretation of the findings. MAP, AF, MK, and RA drafted the first version of the manuscript. All authors critically revised the manuscript for important intellectual content. All authors accept full responsibility for the content of the paper and have seen and approved the final version. AF, MK, and RA contributed equally to this study.

## Data sharing

Deidentified individual participant data underlying the findings of this study will be available for researchers submitting a methodologically sound proposal to achieve the aims of the proposal after publication of this article. Proposals should be directed at the corresponding author (Prof. Dr. Milo A. Puhan).

## Acknowledgements

This study is part of the Corona Immunitas research network, coordinated by the Swiss School of Public Health (SSPH+), and funded by fundraising of SSPH+ including funds of the Swiss Federal Office of Public Health and private funders (ethical guidelines for funding stated by SSPH+ were respected), by funds of the cantons of Switzerland (Vaud, Zurich, and Basel), and by institutional funds of the Universities.

The authors thank the study administration teams in Ticino, Vaud, and Zurich for their dedicated support of the study and the study participants for their valuable contribution to this project.

## CORONA IMMUNITAS RESEARCH GROUP (25 March 2022

**Emiliano Albanese**, MD, PhD (Institute of Public Health (IPH), Università della Svizzera italiana, Lugano, Switzerland); **Rebecca Amati**, PhD (Institute of Public Health (IPH), Università della Svizzera italiana, Lugano, Switzerland) **Antonio Amendola**, Msc (Department of Business Economics, Health and Social Care (DEASS),, University of Applied Sciences & Arts of Southern Switzerland (SUPSI), Switzerland; **Alexia Anagnostopoulos**, MD MPH (Epidemiology, Biostatistics and Prevention Institute, University of Zurich, Zurich, Switzerland); **Daniela Anker**, PhD (Population Health Laboratory (#PopHealthLab), University of Fribourg, Switzerland); Institute of Primary Health Care (BIHAM), University of Bern, Switzerland); **Anna Maria Annoni**, Msc (Institute of Public Health (IPH), Università della Svizzera italiana, Lugano, Switzerland); **Hélène Aschmann**, PhD (Epidemiology, Biostatistics and Prevention Institute, University of Zurich, Zurich, Switzerland); **Andrew Azman**, PhD (Unit of Population Epidemiology, Division of Primary Care Medicine, Geneva University Hospitals, Geneva, Switzerland; Department of Epidemiology, Johns Hopkins Bloomberg School of Public Health, Baltimore, MD, USA; Institute of Global Health, Faculty of Medicine, University of Geneva, Geneva, Switzerland); **Antoine Bal**, MSc (Unit of Population Epidemiology, Division of Primary Care Medicine, Geneva University Hospitals, Geneva, Switzerland); **Tala Ballouz**, MD MPH (Epidemiology, Biostatistics and Prevention Institute, University of Zurich, Zurich, Switzerland); **Hélène Baysson**, PhD (Unit of Population Epidemiology, Division of Primary Care Medicine, Geneva University Hospitals, Geneva, Switzerland; Department of Health and Community Medicine, Faculty of Medicine, University of Geneva, Geneva, Switzerland); **Kleona Bezani**, Msc (Institute of Public Health (IPH), Università della Svizzera italiana, Lugano, Switzerland); **Annette Blattmann** (Cantonal Hospital St. Gallen, Clinic for Infectious Diseases and Hospital Epidemiology, St. Gallen, Switzerland); **Patrick Bleich** (Unit of Population Epidemiology, Division of Primary Care Medicine, Geneva University Hospitals, Geneva, Switzerland); **Murielle Bochud**, MD, PhD (Center for Primary Care and Public Health (Unisanté), University of Lausanne, Switzerland); **Patrick Bodenmann**, MD, Msc (Center for Primary Care and Public Health (Unisanté), University of Lausanne, Switzerland); **Gaëlle Bryand Rumley**, MSc (Unit of Population Epidemiology, Division of Primary Care Medicine, Geneva University Hospitals, Geneva, Switzerland); **Peter Buttaroni** (Institute of Public Health (IPH), Università della Svizzera italiana, Lugano, Switzerland); **Audrey Butty**, MD (Center for Primary Care and Public Health (Unisanté), University of Lausanne, Switzerland); **Anne Linda Camerini**, PhD (Institute of Public Health (IPH), Università della Svizzera italiana, Lugano, Switzerland); **Arnaud Chiolero**, MD, PhD (Population Health Laboratory (#PopHealthLab), University of Fribourg, Switzerland; Institute of Primary Health Care (BIHAM), University of Bern, Switzerland; Department of Epidemiology, Biostatistics and Occupational Health, McGill University, Montréal, Canada); **Patricia Orializ Chocano-Bedoya**, MD, PhD (Institute of Primary Health Care (BIHAM), University of Bern; Population Health Laboratory (#PopHealthLab), University of Fribourg, Switzerland); **Prune Collombet** (Unit of Population Epidemiology, Division of Primary Care Medicine, Geneva University Hospitals, Geneva, Switzerland; Department of Health and Community Medicine, Faculty of Medicine, University of Geneva, Geneva, Switzerland); **Laurie Corna**, PhD (Department of Business Economics, Health and Social Care (DEASS),, University of Applied Sciences & Arts of Southern Switzerland (SUPSI), Switzerland); **Luca Crivelli**, PhD (Department of Business Economics, Health and Social Care (DEASS), University of Applied Sciences & Arts of Southern Switzerland (SUPSI), Switzerland); Institute of Public Health (IPH), Università della Svizzera italiana, Lugano, Switzerland); **Stéphane Cullati**, PhD (Population Health Laboratory (#PopHealthLab), University of Fribourg, Switzerland; Department of Readaptation and Geriatrics, University of Geneva, Switzerland); **Valérie D’Acremont**, MD, PhD (Center for Primary Care and Public Health (Unisanté), University of Lausanne, Switzerland; Swiss Tropical and Public Health Institute, Basel, Switzerland); **Diana Sofia Da Costa Santos** (Institute of Public Health (IPH), Università della Svizzera italiana, Lugano, Switzerland); **Agathe Deschamps** (Cantonal Medical Service Neuchâtel); **Paola D’Ippolito** (Unit of Population Epidemiology, Division of Primary Care Medicine, Geneva University Hospitals, Geneva, Switzerland); **Anja Domenghino**, Dr. med. (Epidemiology, Biostatistics and Prevention Institute, University of Zurich, Zurich, Switzerland); **Richard Dubos**, MSc (Unit of Population Epidemiology, Division of Primary Care Medicine, Geneva University Hospitals, Geneva, Switzerland); **Roxane Dumont**, MSc (Unit of Population Epidemiology, Division of Primary Care Medicine, Geneva University Hospitals, Geneva, Switzerland); **Olivier Duperrex**, MD,MSc (Center for Primary Care and Public Health (Unisanté), University of Lausanne, Switzerland); **Julien Dupraz**, MD, MAS (Center for Primary Care and Public Health (Unisanté), University of Lausanne, Switzerland); **Malik Egger** (Center for Primary Care and Public Health (Unisanté), University of Lausanne, Switzerland); **Emna El-May**, MSc (Population Health Laboratory (#PopHealthLab), University of Fribourg, Switzerland); **Nacira El Merjani** (Unit of Population Epidemiology, Division of Primary Care Medicine, Geneva University Hospitals, Geneva, Switzerland); **Nathalie Engler** (Cantonal Hospital St. Gallen, Clinic for Infectious Diseases and Hospital Epidemiology, St. Gallen, Switzerland); **Adina Mihaela Epure**, MD (Population Health Laboratory (#PopHealthLab), University of Fribourg, Switzerland); **Lukas Erksam** (Institute of Primary Health Care (BIHAM), University of Bern, Department of General Internal Medicine, Inselspital, Bern University Hospital, University of Bern); **Sandrine Estoppey** (Center for Primary Care and Public Health (Unisanté), University of Lausanne, Switzerland); **Marta Fadda**, PhD (Institute of Public Health (IPH), Università della Svizzera italiana, Lugano, Switzerland); **Vincent Faivre** (Center for Primary Care and Public Health (Unisanté), University of Lausanne, Switzerland); **Jan Fehr**, MD (Epidemiology, Biostatistics and Prevention Institute, University of Zurich, Zurich, Switzerland); **Andrea Felappi** (Center for Primary Care and Public Health (Unisanté), University of Lausanne, Switzerland); **Maddalena Fiordelli**, PhD (Institute of Public Health (IPH), Università della Svizzera italiana, Lugano, Switzerland); **Antoine Flahault**, MD, PhD (Institute of Global Health, Faculty of Medicine, University of Geneva, Geneva, Switzerland; Division of Tropical and Humanitarian Medicine, Geneva University Hospitals, Geneva, Switzerland; Department of Health and Community Medicine, Faculty of Medicine, University of Geneva, Geneva, Switzerland); **Luc Fornerod**, MAS (Observatoire valaisan de la santé (OVS), Sion, Switzerland); **Cristina Fragoso Corti**, PhD (Department of environment construction and design (DACD, University of Applied Sciences & Arts of Southern Switzerland (SUPSI), Switzerland); **Natalie Francioli** (Unit of Population Epidemiology, Division of Primary Care Medicine, Geneva University Hospitals, Geneva, Switzerland); **Marion Frangville**, MSc (Unit of Population Epidemiology, Division of Primary Care Medicine, Geneva University Hospitals, Geneva, Switzerland); **Irène Frank**, PhD (Luzerner Kantonsspital, Spitalstrasse, 6000 Luzern 16); **Giovanni Franscella**, Msc (Institute of Public Health (IPH), Università della Svizzera italiana, Lugano, Switzerland**); Anja Frei**, PhD (Epidemiology, Biostatistics and Prevention Institute, University of Zurich, Zurich, Switzerland); **Marco Geigges**, PhD (Epidemiology, Biostatistics and Prevention Institute, University of Zurich, Zurich, Switzerland); **Semira Gonseth Nusslé**, MD, MSc (Center for Primary Care and Public Health (Unisanté), University of Lausanne, Switzerland); **Clément Graindorge**, MD (Unit of Population Epidemiology, Division of Primary Care Medicine, Geneva University Hospitals, Geneva, Switzerland); **Idris Guessous**, MD, PhD (Unit of Population Epidemiology, Division of Primary Care Medicine, Geneva University Hospitals, Geneva, Switzerland; Department of Health and Community Medicine, Faculty of Medicine, University of Geneva, Geneva, Switzerland); **Erika Harju**, PhD (Department of Health Sciences and Medicine, University of Lucerne, Frohburgstrasse 3, 6002 Lucerne); **Séverine Harnal** (Unit of Population Epidemiology, Division of Primary Care Medicine, Geneva University Hospitals, Geneva, Switzerland); **Medea Imboden**, PhD (Swiss Tropical and Public Health Institute, Department of Epidemiology and Public Health, Basel, Switzerland; University of Basel, Basel, Switzerland); **Emilie Jendly** (Center for Primary Care and Public Health (Unisanté), University of Lausanne, Switzerland); **Ayoung Jeong**, PhD (Swiss Tropical and Public Health Institute, Department of Epidemiology and Public Health, Basel, Switzerland; University of Basel, Basel, Switzerland); **Christian R Kahlert**, MD (Cantonal Hospital St. Gallen, Clinic for Infectious Diseases and Hospital Epidemiology, St. Gallen, Switzerland; Children’s Hospital of Eastern Switzerland, Infectious Diseases and Hospital Epidemiology, St. Gallen, Switzerland); **Laurent Kaiser**, MD, PhD (Geneva Center for Emerging Viral Diseases and Laboratory of Virology, Geneva University Hospitals, Geneva, Switzerland; Division of Infectious Diseases, Geneva University Hospitals, Geneva, Switzerland; Department of Medicine, Faculty of Medicine, University of Geneva, Geneva, Switzerland); **Laurent Kaufmann** (Service de La Santé Publique, Canton de Neuchâtel, Neuchâtel, Switzerland); **Marco Kaufmann** PhD (Epidemiology, Biostatistics and Prevention Institute, University of Zurich, Zurich, Switzerland); **Dirk Keidel**, MSc (Swiss Tropical and Public Health Institute, Department of Epidemiology and Public Health, Basel, Switzerland; University of Basel, Basel, Switzerland); **Simone Kessler** (Cantonal Hospital St. Gallen, Clinic for Infectious Diseases and Hospital Epidemiology, St. Gallen, Switzerland); **Philipp Kohler**, MD, MPH (Cantonal Hospital St. Gallen, Clinic for Infectious Diseases and Hospital Epidemiology, St. Gallen, Switzerland); **Christine Krähenbühl** (Luzerner Kantonsspital, Spitalstrasse, 6000 Luzern 16); **Susi Kriemler**, MD (Epidemiology, Biostatistics and Prevention Institute, University of Zurich, Zurich, Switzerland); **Julien Lamour** (Unit of Population Epidemiology, Division of Primary Care Medicine, Geneva University Hospitals, Geneva, Switzerland); **Sara Levati**, PhD (Department of Business Economics, Health and Social Care (DEASS), University of Applied Sciences & Arts of Southern Switzerland (SUPSI), Switzerland); **Pierre Lescuyer**, PhD (Division of Laboratory Medicine, Geneva University Hospitals, Geneva, Switzerland); **Andrea Loizeau**, PhD (Unit of Population Epidemiology, Division of Primary Care Medicine, Geneva University Hospitals, Geneva, Switzerland); **Elsa Lorthe**, RM, PhD (Unit of Population Epidemiology, Division of Primary Care Medicine, Geneva University Hospitals, Geneva, Switzerland); **Chantal Luedi** (Department Health Sciences and Medicine, University of Lucerne, Frohburgstrasse 3, 6002 Lucerne); **Jean-Luc Magnin**, PhD (Laboratory, HFR-Fribourg, Fribourg, Switzerland); **Chantal Martinez** (Unit of Population Epidemiology, Division of Primary Care Medicine, Geneva University Hospitals, Geneva, Switzerland); **Eric Masserey** (Cantonal Medical Office, General Health Department, Canton of Vaud, Switzerland); **Dominik Menges**, MD MPH (Epidemiology, Biostatistics and Prevention Institute, University of Zurich, Zurich, Switzerland); **Gisela Michel**, PhD (Department of Health Sciences and Medicine, University of Lucerne, Frohburgstrasse 3, 6002 Lucerne); **Rosalba Morese**, PhD (Faculty of Communication, Culture and Society, Università della Svizzera italiana, Lugano, Switzerland; Faculty of Biomedical Sciences, Università della Svizzera italiana, Lugano, Switzerland); **Nicolai Mösli** (Swiss TPH, Basel, Switzerland; University of Basel, Basel, Swtizerland); **Natacha Noël** (Unit of Population Epidemiology, Division of Primary Care Medicine, Geneva University Hospitals, Geneva, Switzerland); **Daniel Henry Paris**, MD PhD (Swiss TPH, Basel, Switzerland; University of Basel, Basel, Swtizerland); **Jérôme Pasquier**, PhD (Center for Primary Care and Public Health (Unisanté), University of Lausanne, Switzerland); **Francesco Pennacchio**, PhD (Unit of Population Epidemiology, Division of Primary Care Medicine, Geneva University Hospitals, Geneva, Switzerland**); Stefan Pfister**, PhD (Laboratory, HFR-Fribourg, Fribourg, Switzerland); **Giovanni Piumatti**, PhD (Fondazione Agnelli, Turin, Italy); **Géraldine Poulain** (Division of Laboratory Medicine, Geneva University Hospitals, Geneva, Switzerland); **Nicole Probst-Hensch**, Dr. phil.II, PhD, MPH (Swiss Tropical and Public Health Institute, Department of Epidemiology and Public Health, Basel, Switzerland; University of Basel, Basel, Swtizerland); **Caroline Pugin** (Unit of Population Epidemiology, Division of Primary Care Medicine, Geneva University Hospitals, Geneva, Switzerland); **Milo Puhan**, MD, PhD (Epidemiology, Biostatistics and Prevention Institute, University of Zurich, Zurich, Switzerland); **Nick Pullen**, PhD (Unit of Population Epidemiology, Division of Primary Care Medicine, Geneva University Hospitals, Geneva, Switzerland); **Thomas Radtke**, PhD (Epidemiology, Biostatistics and Prevention Institute, University of Zurich, Zurich, Switzerland); **Manuela Rasi**, MScN (Epidemiology, Biostatistics and Prevention Institute, University of Zurich, Zurich, Switzerland); **Aude Richard** (Unit of Population Epidemiology, Division of Primary Care Medicine, Geneva University Hospitals, Geneva, Switzerland; Institute of Global Health, University of Geneva, Switzerland); **Viviane Richard**, MSc (Unit of Population Epidemiology, Division of Primary Care Medicine, Geneva University Hospitals, Geneva, Switzerland); **Claude-François Robert** (Cantonal Medical Service Neuchâtel); **Pierre-Yves Rodondi**, MD (Institute of Family Medicine, University of Fribourg, Fribourg, Switzerland); **Nicolas Rodondi**, MD, MAS (Institute of Primary Health Care (BIHAM), University of Bern; Department of General Internal Medicine, Inselspital, Bern University Hospital, University of Bern); **Serena Sabatini**, PhD (Institute of Public Health (IPH), Università della Svizzera italiana, Lugano, Switzerland); **Khadija Samir** (Unit of Population Epidemiology, Division of Primary Care Medicine, Geneva University Hospitals, Geneva, Switzerland); **Javier Sanchis Zozaya**, MD (Center for Primary Care and Public Health (Unisanté), University of Lausanne, Switzerland); **Virginie Schlüter**, MAS (Center for Primary Care and Public Health (Unisanté), University of Lausanne, Switzerland); **Alexia Schmid**, MSc (Institute of Family Medicine, University of Fribourg, Fribourg, Switzerland); **Valentine Schneider** (Cantonal Medical Service Neuchâtel); **Maria Schüpbach** (Institute of Primary Health Care (BIHAM), University of Bern, Department of General Internal Medicine, Inselspital, Bern University Hospital, University of Bern); **Nathalie Schwab** (Institute of Primary Health Care (BIHAM), University of Bern, Department of General Internal Medicine, Inselspital, Bern University Hospital, University of Bern);); **Claire Semaani** (Unit of Population Epidemiology, Division of Primary Care Medicine, Geneva University Hospitals, Geneva, Switzerland); **Alexandre Speierer** (Institute of Primary Health Care (BIHAM), University of Bern; Department of General Internal Medicine, Inselspital, Bern University Hospital, University of Bern); **Amélie Steiner-Dubuis** (Center for Primary Care and Public Health (Unisanté), University of Lausanne, Switzerland); **Silvia Stringhini**, PhD (Unit of Population Epidemiology, Division of Primary Care Medicine, Geneva University Hospitals, Geneva, Switzerland; Department of Health and Community Medicine, Faculty of Medicine, University of Geneva, Geneva, Switzerland); **Stefano Tancredi**, MD (Population Health Laboratory (#PopHealthLab), University of Fribourg, Switzerland); **Stéphanie Testini** (Unit of Population Epidemiology, Division of Primary Care Medicine, Geneva University Hospitals, Geneva, Switzerland); **Julien Thabard** (Center for Primary Care and Public Health (Unisanté), University of Lausanne, Switzerland); **Mauro Tonolla**, PD PhD (Department of environment construction and design (DACD, University of Applied Sciences & Arts of Southern Switzerland (SUPSI), Switzerland); **Nicolas Troillet**, MD, MSc (Office du médecin cantonal, Sion, Switzerland); **Agne Ulyte**, MD (Epidemiology, Biostatistics and Prevention Institute, University of Zurich, Zurich, Switzerland); **Sophie Vassaux** (Center for Primary Care and Public Health (Unisanté), University of Lausanne, Switzerland); **Thomas Vermes**, MSc (Swiss Tropical and Public Health Institute, Department of Epidemiology and Public Health, Basel, Switzerland; University of Basel, Basel, Swtizerland); **Jennifer Villers**, PhD (Unit of Population Epidemiology, Division of Primary Care Medicine, Geneva University Hospitals, Geneva, Switzerland); **Viktor von Wyl** (Epidemiology, Biostatistics and Prevention Institute, University of Zurich, Zurich, Switzerland); **Cornelia Wagner**, MSc (Population Health Laboratory (#PopHealthLab), University of Fribourg, Switzerland); **Rylana Wenger** (Institute of Primary Health Care (BIHAM), University of Bern, Department of General Internal Medicine, Inselspital, Bern University Hospital, University of Bern); **Erin West**, PhD (Epidemiology, Biostatistics and Prevention Institute, University of Zurich, Zurich, Switzerland); **Ania Wisniak**, MD (Unit of Population Epidemiology, Division of Primary Care Medicine, Geneva University Hospitals, Geneva, Switzerland; Institute of Global Health, Faculty of Medicine, University of Geneva, Geneva, Switzerland); **Melissa Witzig**, Msc (Swiss Tropical and Public Health Institute, Department of Epidemiology and Public Health, Basel, Switzerland; University of Basel, Basel, Swtizerland); **María-Eugenia Zaballa**, PhD (Unit of Population Epidemiology, Division of Primary Care Medicine, Geneva University Hospitals, Geneva, Switzerland); **Kyra Zens**, PhD, MPH (Epidemiology, Biostatistics and Prevention Institute, University of Zurich, Zurich, Switzerland); **Claire Zuppinger** (Center for Primary Care and Public Health (Unisanté), University of Lausanne, Switzerland)

