## Supplementary Tables S1 and S2 for "Development of hybrid immunity during a period of high incidence of infections with Omicron subvariants: A prospective population based multi-region cohort study"

Supplementary Material

**Supplementary Table S1: Trajectories from March 2022 to June/July 2022 of SARS-CoV-2 IgG antibodies and ACE2r-blocking (neutralising capacity) as measured by a virus-free assay, in participants from Ticino and Zurich, Switzerland, June/July 2022, stratified by age group (n=1702*).**

|  | **Ticino** | **Ticino** | **Ticino** | **Ticino** | **Ticino** | **Zurich** | **Zurich** | **Zurich** | **Zurich** | **Zurich** |
| --- | --- | --- | --- | --- | --- | --- | --- | --- | --- | --- |
| **Age category** | **All** | **16-29** | **30-44** | **45-64** | **65+** | **All** | **16-29** | **30-44** | **45-64** | **65+** |
| **Anti-spike IgG antibodies**** | | |  |  |  |  |  |  |  |  |
| MFI change - Median (IQR) | 1910 (675 to 3574) | 2198 (733 to 4106) | 1641 (727 to 3351) | 2035 (723 to 3919) | 1821 (569 to 2726) | 1686 (503 to 3044) | 1640 (628 to 3065) | 1611 (338 to 3017) | 1669 (655 to 2920) | 1803 (440 to 3199) |
| Negative - Negative | 15 (2%) | 1 (0.8%) | 3 (1.6%) | 7 (2.9%) | 4 (2.2%) | 8 (0.8%) | 1 (0.6%) | 2 (0.9%) | 2 (0.7%) | 3 (1.1%) |
| Negative - Positive | 7 (0.9%) | 2 (1.6%) | 2 (1.1%) | 1 (0.4%) | 2 (1.1%) | 8 (0.8%) | 2 (1.3%) | 1 (0.4%) | 3 (1%) | 2 (0.7%) |
| Positive - Negative | 3 (0.4%) | 3 (2.3%) | 0 (0%) | 0 (0%) | 0 (0%) | 4 (0.4%) | 1 (0.6%) | 3 (1.3%) | 0 (0%) | 0 (0%) |
| Positive - Positive | 713 (96.6%) | 122 (95.3%) | 181 (97.3%) | 233 (96.7%) | 177 (96.7%) | 944 (97.9%) | 152 (97.4%) | 229 (97.4%) | 301 (98.4%) | 262 (98.1%) |
| **Anti-NuC IgG antibodies#** | | |  |  |  |  |  |  |  |  |
| Negative - Negative | 428 (58%) | 63 (49.2%) | 97 (52.2%) | 148 (61.4%) | 120 (65.6%) | 543 (56.3%) | 75 (48.1%) | 114 (48.5%) | 167 (54.6%) | 187 (70%) |
| Negative - Positive | 114 (15.4%) | 19 (14.8%) | 29 (15.6%) | 35 (14.5%) | 31 (16.9%) | 202 (21%) | 35 (22.4%) | 53 (22.6%) | 64 (20.9%) | 50 (18.7%) |
| Positive - Negative | 56 (7.6%) | 18 (14.1%) | 18 (9.7%) | 12 (5%) | 8 (4.4%) | 69 (7.2%) | 15 (9.6%) | 20 (8.5%) | 28 (9.2%) | 6 (2.2%) |
| Positive - Positive | 140 (19%) | 28 (21.9%) | 42 (22.6%) | 46 (19.1%) | 24 (13.1%) | 150 (15.6%) | 31 (19.9%) | 48 (20.4%) | 47 (15.4%) | 24 (9%) |
| **Neutralisation wildtype##** | | |  |  |  |  |  |  |  |  |
| Negative - Negative | 43 (5.8%) | 12 (9.4%) | 11 (5.9%) | 13 (5.4%) | 7 (3.8%) | 32 (3.3%) | 5 (3.2%) | 9 (3.8%) | 10 (3.3%) | 8 (3%) |
| Negative - Positive | 10 (1.4%) | 1 (0.8%) | 4 (2.2%) | 4 (1.7%) | 1 (0.5%) | 12 (1.2%) | 2 (1.3%) | 3 (1.3%) | 4 (1.3%) | 3 (1.1%) |
| Positive - Negative | 8 (1.1%) | 2 (1.6%) | 2 (1.1%) | 1 (0.4%) | 3 (1.6%) | 12 (1.2%) | 1 (0.6%) | 4 (1.7%) | 4 (1.3%) | 3 (1.1%) |
| Positive - Positive | 677 (91.7%) | 113 (88.3%) | 169 (90.9%) | 223 (92.5%) | 172 (94%) | 908 (94.2%) | 148 (94.9%) | 219 (93.2%) | 288 (94.1%) | 253 (94.8%) |
| **Neutralisation Delta##** | | |  |  |  |  |  |  |  |  |
| Negative - Negative | 49 (6.6%) | 13 (10.2%) | 12 (6.5%) | 15 (6.2%) | 9 (4.9%) | 45 (4.7%) | 6 (3.8%) | 10 (4.3%) | 13 (4.2%) | 16 (6%) |
| Negative - Positive | 15 (2%) | 2 (1.6%) | 4 (2.2%) | 5 (2.1%) | 4 (2.2%) | 15 (1.6%) | 2 (1.3%) | 3 (1.3%) | 6 (2%) | 4 (1.5%) |
| Positive - Negative | 20 (2.7%) | 5 (3.9%) | 5 (2.7%) | 5 (2.1%) | 5 (2.7%) | 51 (5.3%) | 2 (1.3%) | 18 (7.7%) | 15 (4.9%) | 16 (6%) |
| Positive - Positive | 654 (88.6%) | 108 (84.4%) | 165 (88.7%) | 216 (89.6%) | 165 (90.2%) | 853 (88.5%) | 146 (93.6%) | 204 (86.8%) | 272 (88.9%) | 231 (86.5%) |
| **Neutralisation Omicron##** | | |  |  |  |  |  |  |  |  |
| Negative - Negative | 62 (8.4%) | 14 (10.9%) | 16 (8.6%) | 16 (6.6%) | 16 (8.7%) | 66 (6.8%) | 7 (4.5%) | 17 (7.2%) | 21 (6.9%) | 21 (7.9%) |
| Negative - Positive | 29 (3.9%) | 2 (1.6%) | 6 (3.2%) | 13 (5.4%) | 8 (4.4%) | 37 (3.8%) | 6 (3.8%) | 9 (3.8%) | 12 (3.9%) | 10 (3.7%) |
| Positive - Negative | 54 (7.3%) | 7 (5.5%) | 13 (7%) | 17 (7.1%) | 17 (9.3%) | 92 (9.5%) | 9 (5.8%) | 24 (10.2%) | 25 (8.2%) | 34 (12.7%) |
| Positive - Positive | 593 (80.4%) | 105 (82%) | 151 (81.2%) | 195 (80.9%) | 142 (77.6%) | 769 (79.8%) | 134 (85.9%) | 185 (78.7%) | 248 (81%) | 202 (75.7%) |

Abbreviations: NuC: Nucleocapsid; WHO U / ml: U/ml according to Elecsys ® Anti-SARS-CoV-2 S; IgG: Immunglobulin G

*For one participant from Ticino no results for phase five (March 2022) is available.

**Unit for levels of anti-spike IgG antibodies is the Mean Fluorescence Intensity as measured by the Luminex binding assay SenASTrIS (Sensitive Anti-SARS-CoV-2 Spike Trimer Immunoglobulin Serological).

#Seropositivity is defined based on the presence of anti-spike IgG antibodies according to the threshold of SenASTrIS test positivity with mean MFI ≥ 6.

##Neutralisation capacity based on the cut-off value of MFI ≥ 50.

**Supplementary Table S2: Prevalence of SARS-CoV-2 IgG antibodies and ACE2r-blocking (neutralising capacity) as measured by a virus-free assay, stratified by vaccination and infection status of participants, Ticino, Vaud, and Zurich, Switzerland, June/July 2022, (n=2520*)**

|  | **Ticino** | **Ticino** | **Ticino** | **Vaud** | **Vaud** | **Vaud** | **Zurich** | **Zurich** | **Zurich** |
| --- | --- | --- | --- | --- | --- | --- | --- | --- | --- |
| **Immune status** | **Only vaccinated** | **Only infected** | **Vaccinated and infected** | **Only vaccinated** | **Only infected** | **Vaccinated and infected** | **Only vaccinated** | **Only infected** | **Vaccinated and infected** |
| **Sample size** | 285 | 61 | 379 | 354 | 66 | 420 | 411 | 54 | 490 |
| **Anti-spike IgG antibodies**** | | |  |  |  |  |  |  |  |
| Not detectable | 0 (0%) | 6 (9.8%) | 0 (0%) | 0 (0%) | 6 (9.1%) | 1 (0.2%) | 0 (0%) | 5 (9.3%) | 0 (0%) |
| Low (≥6, <12) | 0 (0%) | 8 (13.1%) | 0 (0%) | 0 (0%) | 13 (19.7%) | 0 (0%) | 1 (0.2%) | 9 (16.7%) | 0 (0%) |
| Middle (≥12, <40) | 3 (1.1%) | 16 (26.2%) | 1 (0.3%) | 4 (1.1%) | 20 (30.3%) | 0 (0%) | 3 (0.7%) | 15 (27.8%) | 0 (0%) |
| High (≥40) | 282 (98.9%) | 31 (50.8%) | 378 (99.7%) | 350 (98.9%) | 27 (40.9%) | 419 (99.8%) | 407 (99%) | 25 (46.3%) | 490 (100%) |
| WHO U / ml (median (IQR)) | 4553 (3709-5989) | 628 (64-2257) | 4781 (4020-6460) | 4244 (3269-5266) | 222 (31-1142) | 4284 (3718-5841) | 4172 (3264-5760) | 269 (42-1486) | 4742 (3623-5726) |
| **Anti-NuC IgG antibodies#** | | |  |  |  |  |  |  |  |
| Not detectable | 285 (100%) | 21 (34.4%) | 166 (43.8%) | 354 (100%) | 22 (33.3%) | 179 (42.6%) | 411 (100%) | 16 (29.6%) | 177 (36.1%) |
| Low (≥6, <12) | 0 (0%) | 12 (19.7%) | 120 (31.7%) | 0 (0%) | 16 (24.2%) | 122 (29%) | 0 (0%) | 10 (18.5%) | 165 (33.7%) |
| Middle (≥12, <40) | 0 (0%) | 28 (45.9%) | 93 (24.5%) | 0 (0%) | 28 (42.4%) | 119 (28.3%) | 0 (0%) | 28 (51.9%) | 148 (30.2%) |
| High (≥40) | 0 (0%) | 0 (0%) | 0 (0%) | 0 (0%) | 0 (0%) | 0 (0%) | 0 (0%) | 0 (0%) | 0 (0%) |
| **Neutralisation (≥50)##** | |  |  |  |  |  |  |  |  |
| Wildtype | 280 (98.2%) | 28 (45.9%) | 378 (99.7%) | 349 (98%) | 31 (47%) | 417 (99.8%) | 402 (97.8%) | 27 (50%) | 490 (100%) |
| Delta | 269 (94.4%) | 26 (42.6%) | 373 (98.4%) | 338 (94.9%) | 25 (37.9%) | 416 (99.5%) | 362 (88.1%) | 19 (35.2%) | 486 (99.2%) |
| Omicron | 225 (78.9%) | 29 (47.5%) | 367 (96.8%) | 303 (85.1%) | 32 (48.5%) | 403 (96.4%) | 308 (74.9%) | 25 (46.3%) | 472 (96.3%) |

Abbreviations: NuC: Nucleocapsid; WHO U / ml: U/ml according to Elecsys ® Anti-SARS-CoV-2 S; IgG: Immunglobulin G

*Participants who were immunologically naïve (n=25) or were missing relevant data to determine their immune status (n = 8) have been excluded.

**Unit for levels of anti-spike IgG antibodies is the Mean Fluorescence Intensity as measured by the Luminex binding assay SenASTrIS (Sensitive Anti-SARS-CoV-2 Spike Trimer Immunoglobulin Serological). Low: From threshold of test positivity to less than 3 standard deviations above this threshold (≥6, <12); moderate: ≥3 standard deviations above positivity threshold but unlikely to provide neutralisation (≥12, <40); high: neutralising capacity likely (≥40).

#Seropositivity is defined based on the presence of anti-spike IgG antibodies according to the threshold of SenASTrIS test positivity with mean MFI ≥ 6.

##Neutralisation capacity based on the cut-off value of MFI ≥ 50.
